# Determinants of overweight and obesity among children between 5 to 11 years in Ecuador: a secondary analysis from the National Health Survey 2018

**DOI:** 10.1101/2023.12.16.23300087

**Authors:** Betzabé Tello, José Ocaña, Paúl García-Zambrano, Betsabé Enríque-Moreira, Iván Dueñas-Espín

## Abstract

**Background:** During the 1990s, global eating habits changed, affecting poorer and middle-income nations, as well as richer countries. This shift, known as the “obesity transition,” led to more people becoming overweight or obese worldwide. In Ecuador, this change is happening, and now, one in three children is affected by overweight or obesity (OW/OB). This study explores the links between social, economic, and demographic factors and childhood obesity in Ecuador, seeking to provide insights for shaping future health policies in response to this intricate shift.

**Methods:** A cross-sectional study using 2018 National Health and Nutrition Survey data from Ecuador. Weighted percentages were computed, and odds ratios for OW/OB unadjusted and adjusted for each category of explanatory variables were estimated using multilevel multivariate logistic regression models.

**Results:** Among 10,807 Ecuadorian school children aged 5 to 11, the prevalence of OW/OB was 36.0%. Males exhibited 1.26 times higher odds than females (95% CI: 1.20 to 1.33), and each additional year of age increased the odds by 1.10 times (95% CI: 1.09 to 1.10). Economic quintiles indicated increased odds (1.17 to 1.39) from the 2nd to 5th quintile (the richest) compared with the first quintile (the poorest). Larger household size slightly reduced odds of OW/OB (adjusted odds ratio [aOR]=0.93, 95% CI: 0.91 to 0.95), while regular physical activity decreased odds ([aOR]=0.79, 95% CI: 0.75 to 0.82). The consumption of school-provided meals showed a non-significant reduction (aOR: 0.93, 95% CI: 0.82 to 1.06). Children from families recognizing and using processed food labels had a higher likelihood of being overweight or obese (aOR=1.14, 95% CI: 1.02 to 1.26).

**Conclusion:** Age, male gender, and higher economic quintile increase OW/OB in Ecuadorian school children. Larger households and physical activity slightly decrease risks. Ecuador needs policies for healthy schools and homes, focusing on health, protection, and good eating habits.

## Introduction

Similar to high-income countries, low- and middle-income countries-initiated processes of nutritional and food environment transition in the 1990s. These transitions are characterized by an increase in the consumption of processed foods, edible oils, and sugary beverages, as well as a greater tendency to eat outside the home and an increased availability of ultra-processed products. All these changes have come at the expense of healthy and traditional diets. Simultaneously, the population in these countries has gradually reduced physical activity and increased sedentary behavior [1–3].

These transitions have led to a gradual increase in overweight and obesity (OW/OB) in all age groups, with a special increase in childhood OW/OB [1–4]. This global increase follows a pattern known as the “obesity transition.” This pattern is characterized by a gradual shift in the burden of OW/OB from high-income to low- and middle-income countries, from wealthy households to poor ones, from urban to rural areas, and from adults to children. This changes affect several countries in Latin America [5].

Childhood obesity is a precursor to adult cardiovascular diseases and cancer [6–8]. Understanding its determinants is vital for making informed public health policy decisions.

This study aims to identify the independent factors associated with obesity and overweight in Ecuadorian school-age children (5-11 years). By delving into obesogenic environments and contextual sociodemographic conditions, this research offers valuable territorial insights.

In Ecuador, the prevalence of childhood OW/OB surged by almost 5 percentage points from 2012 to 2018, reaching 35.4% [9]. This alarming increase predominantly impacts urban areas, males, those with mixed and white ethnic backgrounds, and wealthier households. Importantly, this trend extends beyond, affecting middle- and low-income households and rural populations, highlighting the urgency for a thorough exploration of its determinants [9].

While there is extensive knowledge about the social, environmental, and clinical determinants of excess malnutrition in school-age children, particularly in high-income countries [10], few countries in the region have studied these determinants [11,12]. In low- and middle-income countries such as Ecuador, there is a lack of national information regarding the determinants of childhood obesity in school-age children, except for some localized studies [13–15]. The existing knowledge gap precludes the development of effective public health policies. Despite the bulk of scientific evidence, the Ecuadorian government’s efforts are not focused on improving school environments, increasing taxes on sugary beverages, or improving the labeling of processed and ultra-processed products; rather, it has reduced taxes on ultra-processed food [16]. Therefore, initiatives with proper implementation, supervision, and robust evaluations are necessary to demonstrate their impact and cost-effectiveness in school-age populations. These insights will serve as a compass for evidence-based public policies and interventions, which are crucial for combating childhood obesity in Ecuador.

## Materials and Methods

### Study design

This cross-sectional study involved a secondary analysis, utilizing data from the 2018 National Health and Nutrition Survey of Ecuador. To ensure methodological rigor and transparency in both the study design and dissemination of findings, compliance with the Strengthening the Reporting of Observational Studies in Epidemiology (STROBE) [17] guidelines was scrupulously maintained, as detailed in S1 Table.

### Population and sample

In our study, we included data from children encompassing sociodemographic information, anthropometric measurements, dietary habits at home and school, and physical activity status. Additionally, we integrated information about whether the children’s households identified, understood, and used the nutritional traffic light labeling system for processed foods and beverages that was implemented in Ecuador. These questions were incorporated into the National Health and Nutrition Survey of 2018 (see flowchart in Fig 1). We included study subjects who: (i) are ≥5 years old or ≤11 years old; (ii) had complete anthropometric information, and (iii) had complete data regarding age, ethnicity, economic quintile, schooling characteristics, dietary habits, and physical activity.

**Figure 1.**
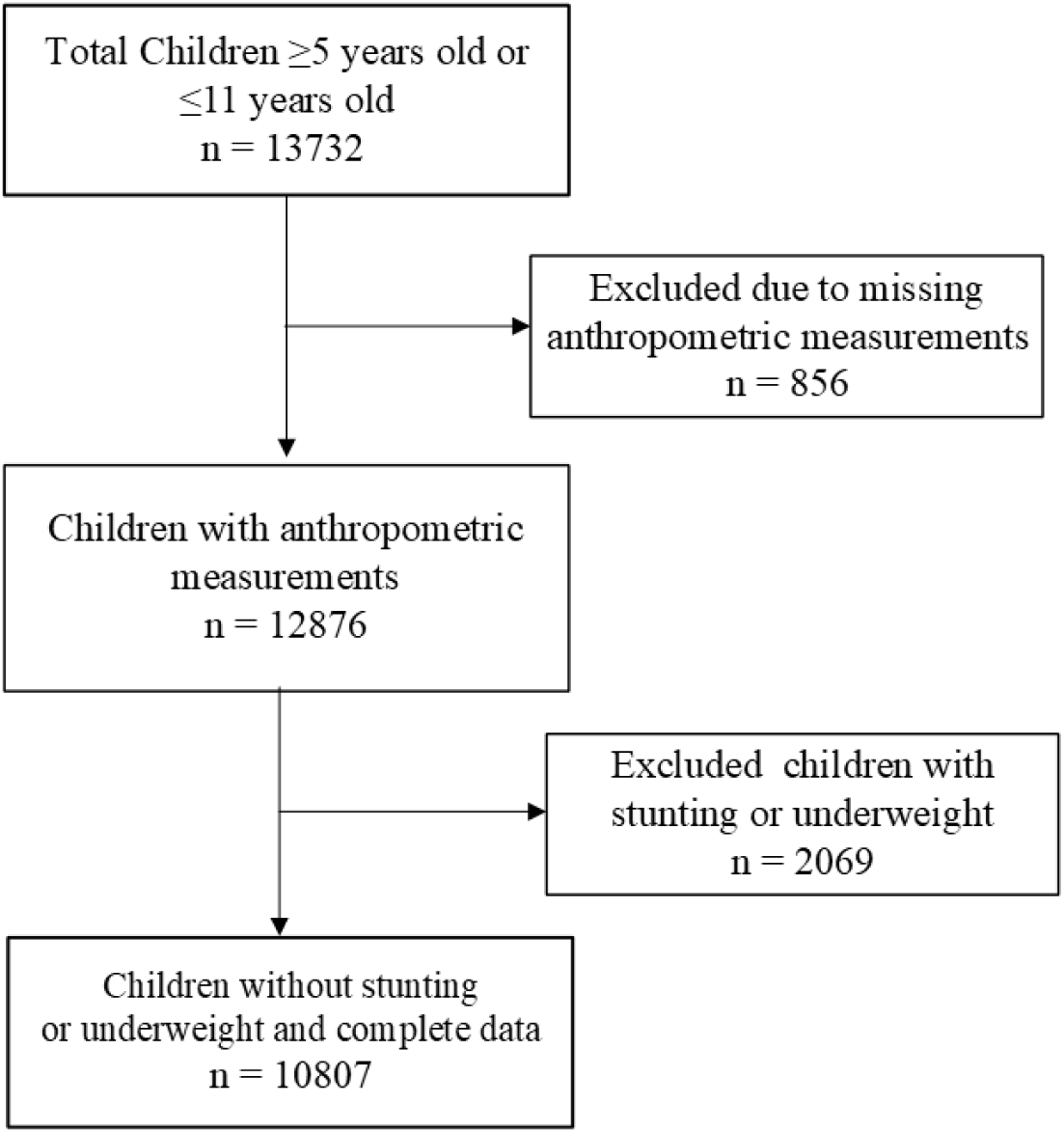
Flowchart showing the study population and selection of study participants

### The ENSANUT 2018 survey

The Ecuadorian National Health and Nutrition Survey 2018 (ENSANUT 2018, for its acronym in Spanish) was a cross-sectional study conducted in 2018 that involved nationally representative samples from the Ecuadorian population [18].

In the ENSANUT 2018 study, a two-stage sampling strategy was employed to secure a representative sample of the Ecuadorian populace. Initially, Primary Sampling Units (PSU) were chosen through stratified sampling, incorporating proportional probability to size. Subsequently, an average of 18 households per PSU were randomly selected for investigation. Within these households, targeted demographic groups were identified, including children under 5 years, women aged 12 to 49, men aged over 12 years, and individuals between 5 and 17 years. For households with children aged 5 to 11 years, one individual between 5 and 17 years was selected for interview and subjected to a specialized questionnaire. Anthropometric measurements were obtained. The abovementioned sampling approach ensured the data quality and representativeness of the study. Further information on the methodology, datasets, and findings of ENSANUT 2018 is available at: https://www.ecuadorencifras.gob.ec/institucional/home/.

### Measurements

We used the information that the survey collected about sex, age of the child, ethnicity, education of the children, economic quintile, regular class attendance, geographical regions of Ecuador, receiving the human developing bonus (BDH, for its acronym in Spanish), number of people in the household, disposal of excreta, physical activity, perception of consumption of vegetables, consumption of fast food, days per week of school food consumption, buying food at school, consumption of the food provided by the school, recognizing, understanding, and using the nutritional traffic light labelling of processed foods; and, consumption of processed foods with a red label. The Ecuadorian Nutritional Traffic Light Labelling system uses three colours - red, yellow, and green - to indicate the levels of sugar, fat, and salt in processed foods. Red signifies high concentrations, yellow denotes medium levels, and green indicates low content [19]. This system empowers consumers to make healthier food choices.

### Main outcome

The main outcome variable was, overweight or obesity. The World Health Organization (WHO) macro program Stata (WHO AnthroPlus) was used to establish the nutritional status of children based on WHO 2007 standards for the classification of children as overweight or obese between 5 and 19 years. Overweight was defined as a Body-Mass-Index (BMI)-for-age greater than 1 standard deviation above the WHO Growth Reference median; and obesity as a greater than 2 standard deviations above the WHO Growth Reference median [20].

### Statistical analyses and sample considerations

A priori, we calculated that a sample of 9759 individuals is enough to estimate, with 95% confidence and an accuracy of +/- 1 percentage units, a population percentage that will predictably be around 35.38% [9]. The percentage of necessary replacements is expected to be 10%.

In the analysis, the ‘svy’ command from Stata 16.1 (StataCorp. 2019. Stata Statistical Software: Release 16. College Station, TX: StataCorp LLC.) was employed to accommodate the expansion factors inherent in the survey design. This methodology yield estimates that faithfully represent the complex nature of the survey’s structure. Descriptive statistics for complex survey data involved calculating weighted percentages for categorical variables and deriving means and standard errors for discrete ones. Subsequently, the characteristics of non-overweight or obese children were compared with those of overweight or obese children, adhering to the same ‘svy’ command approach.

Then, multilevel logistic regression models were built to analyze the relationships between the explanatory variables and the outcome. We estimated crude and adjusted odds ratios (OR and aOR) of OW/OB for each explanatory variable and/or their categories. In that sense, we built multilevel multivariate logistic regression models to evaluate the independent association between each explanatory variable and health self-perception. The multilevel variable was geographical region, considering that there are important differences in OW/OB across those regions. Moreover, expansion factors were used to align estimates with the stratified sampling method and primary sampling units, managing variability and correlations within groups.

We built a saturated model that included all the individual covariates. Then, based on the researchers’ criteria, we eliminated covariates with p>0.25 from significant covariates that were retained in the model [21]. Confidence intervals (95%CI) of the adjusted and unadjusted ORs and their corresponding p-values were calculated. Once the parsimonious model was obtained, we compared both models and chose the “final” model, according to its level of significance from the likelihood ratio test. The final model was stratified by sex. Given the small number of missing data (there were missing values in <1% of the whole database), we employed complete case analysis to estimate statistical associations.

To test for potential effect modification, we performed several secondary analyses to assess the sensitivity of our estimates with our assumptions regarding biases, and to test for model misspecifications. We ran the final model excluding: (i) children categorized in the highest income quintile, (ii) children whose parents received the BDH, and (iii) children within the upper third of the highest number of people per household.

### Ethical issues

The research protocol was thoroughly reviewed and approved by the Ethics Subcommittee for Research in Human Beings of the Faculty of Medicine of the Pontifical Catholic University of Ecuador under the code SB-CEISH-POS-691. The committee determined that informed consent was not required for this study.

## Results

A comprehensive demographic and health snapshot of children aged 5 to 11 years is presented in Table 1. In the study sample comprising 10807 children aged 5 to 11 from Ecuador, it was found that the mean age of the children was 8.0 years (standard error [SE]: 0.03). About ethnicity, Mestizo (mixed ethnic background) children were predominant, making up 80.8% of the sample. When stratified by economic quintiles, approximately a quarter of the children (25.4%) belonged to the lowest income quintile. Educationally, it was noted that the vast majority (98.3%) were currently attending elementary school. Regarding dietary habits, a significant proportion of the sample (64.3%) reported consuming food provided by the school, and nearly half (48.4%) indicated a reduction in the consumption of processed foods with a red label. Notably, 36.0% of the children were identified as being overweight or obese, according to the Body Mass Index (BMI) adjusted for age and sex, using the WHO Growth References.

**Table 1.**
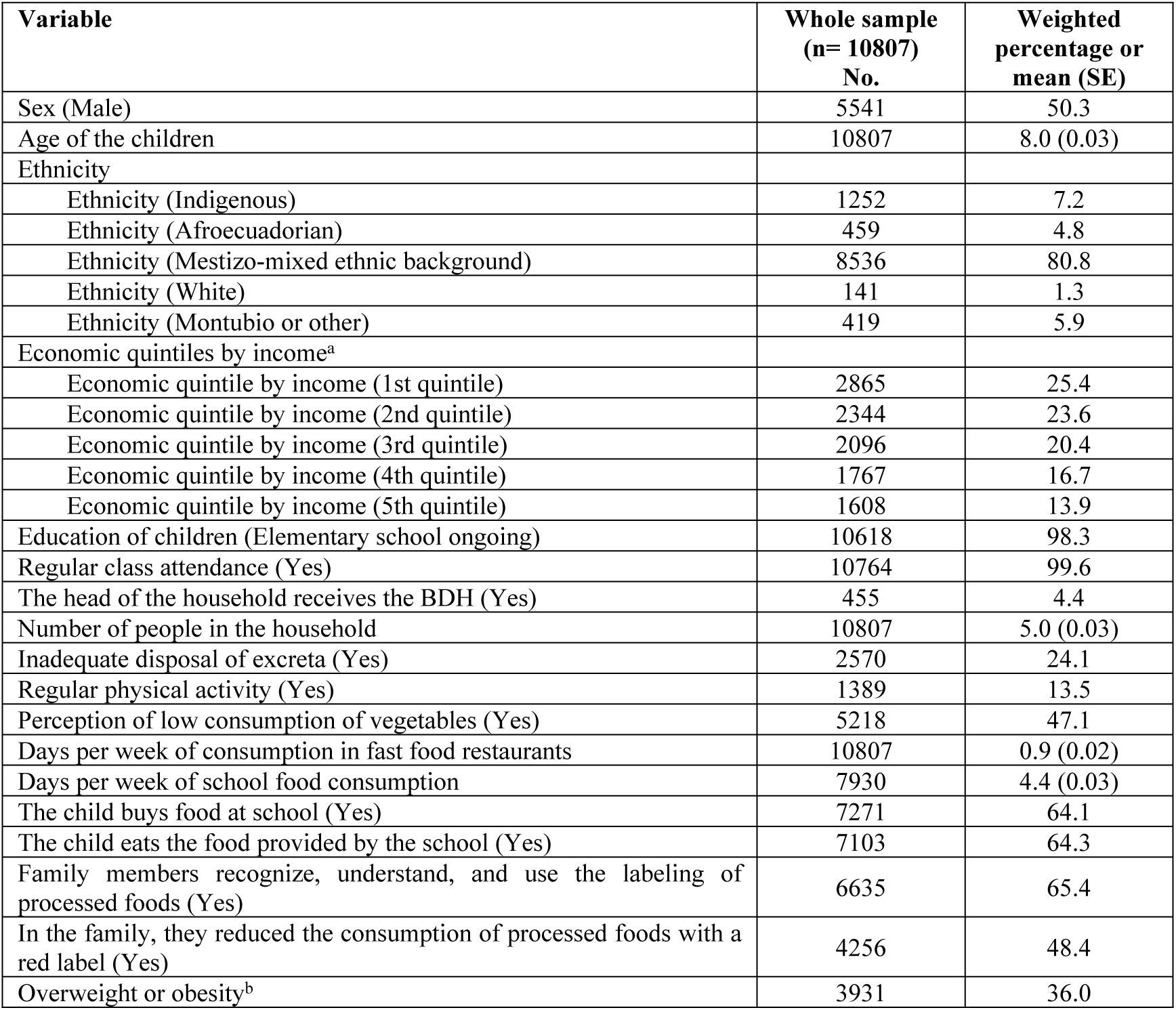
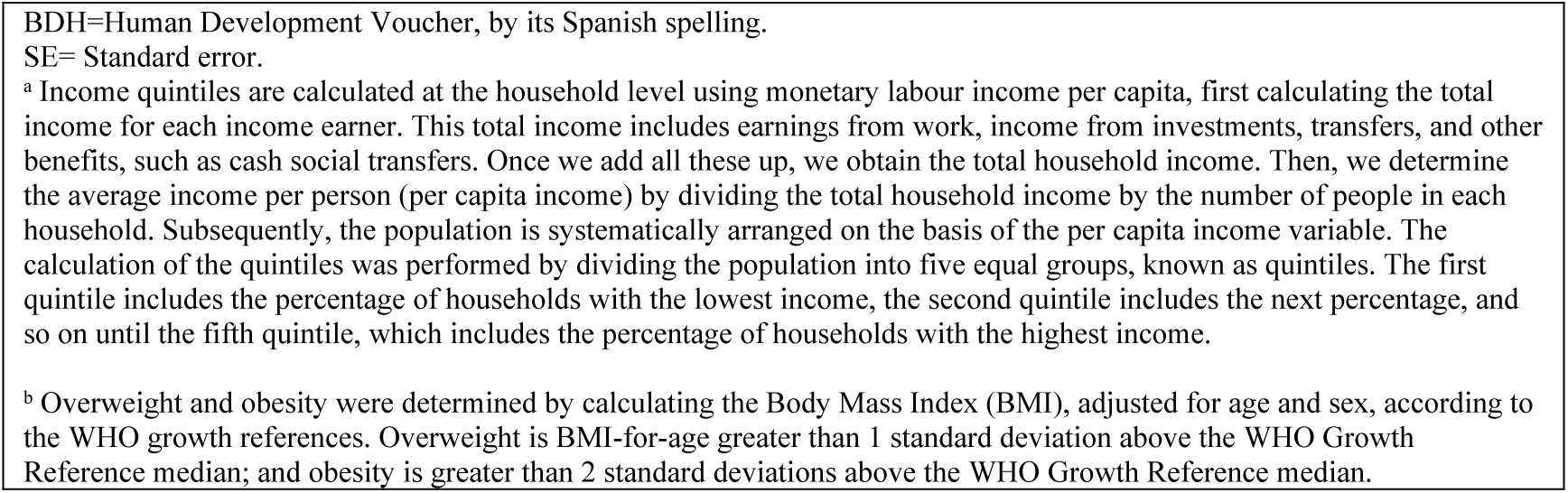
Description of the sample.

| Variable | Whole sample<br>(n= 10807)<br>No. | Weighted<br>percentage or<br>mean (SE) |
| --- | --- | --- |
| Sex (Male) | 5541 | 50.3 |
| Age of the children | 10807 | 8.0 (0.03) |
| Ethnicity |  |  |
| Ethnicity (Indigenous) | 1252 | 7.2 |
| Ethnicity (Afroecuadorian) | 459 | 4.8 |
| Ethnicity (Mestizo-mixed ethnic background) | 8536 | 80.8 |
| Ethnicity (White) | 141 | 1.3 |
| Ethnicity (Montubio or other) | 419 | 5.9 |
| Economic quintiles by income<sup>a</sup> |  |  |
| Economic quintile by income (1st quintile) | 2865 | 25.4 |
| Economic quintile by income (2nd quintile) | 2344 | 23.6 |
| Economic quintile by income (3rd quintile) | 2096 | 20.4 |
| Economic quintile by income (4th quintile) | 1767 | 16.7 |
| Economic quintile by income (5th quintile) | 1608 | 13.9 |
| Education of children (Elementary school ongoing) | 10618 | 98.3 |
| Regular class attendance (Yes) | 10764 | 99.6 |
| The head of the household receives the BDH (Yes) | 455 | 4.4 |
| Number of people in the household | 10807 | 5.0 (0.03) |
| Inadequate disposal of excreta (Yes) | 2570 | 24.1 |
| Regular physical activity (Yes) | 1389 | 13.5 |
| Perception of low consumption of vegetables (Yes) | 5218 | 47.1 |
| Days per week of consumption in fast food restaurants | 10807 | 0.9 (0.02) |
| Days per week of school food consumption | 7930 | 4.4 (0.03) |
| The child buys food at school (Yes) | 7271 | 64.1 |
| The child eats the food provided by the school (Yes) | 7103 | 64.3 |
| Family members recognize, understand, and use the labeling of processed foods (Yes) | 6635 | 65.4 |
| In the family, they reduced the consumption of processed foods with a red label (Yes) | 4256 | 48.4 |
| Overweight or obesity<sup>b</sup> | 3931 | 36.0 |

BDH=Human Development Voucher, by its Spanish spelling.
SE= Standard error.
<sup>a</sup> Income quintiles are calculated at the household level using monetary labour income per capita, first calculating the total income for each income earner. This total income includes earnings from work, income from investments, transfers, and other benefits, such as cash social transfers. Once we add all these up, we obtain the total household income. Then, we determine the average income per person (per capita income) by dividing the total household income by the number of people in each household. Subsequently, the population is systematically arranged on the basis of the per capita income variable. The calculation of the quintiles was performed by dividing the population into five equal groups, known as quintiles. The first quintile includes the percentage of households with the lowest income, the second quintile includes the next percentage, and so on until the fifth quintile, which includes the percentage of households with the highest income.
<sup>b</sup> Overweight and obesity were determined by calculating the Body Mass Index (BMI), adjusted for age and sex, according to the WHO growth references. Overweight is BMI-for-age greater than 1 standard deviation above the WHO Growth Reference median; and obesity is greater than 2 standard deviations above the WHO Growth Reference median.

We found distinct differences between non-overweight or non-obese children and their overweight or obese counterparts (S2 Table). In a comparison of characteristics between non-overweight/non-obese children (n=6876) and those identified as overweight or obese (n=3931) the mean age of overweight or obese children was slightly higher at 8.3 years (SE: 0.05) compared to 7.9 years (SE: 0.04) for their non-overweight counterparts. Ethnic distribution showed that 81.7% of the overweight or obese groups were Mestizo (mixed ethnic background), compared with 80.3% in the non-overweight group. Economic stratification revealed that children from the lowest income quintile were less represented (20.6%) among the overweight or obese compared to those non-overweight (28.2%). Furthermore, a greater proportion of overweight or obese children (65.7%) consumed food provided by the school, in contrast to 63.2% of the non-overweight children. Finally, a difference was detected in the perception of reduced consumption of processed foods with a red label, with 50.4% of overweight or obese children indicating consumption, compared to 47.2% of their non-overweight peers.

After running multivariate logistic regression models, we found that, several factors were significantly associated with childhood overweight and obesity (Table 2). According to the final adjusted model, several variables exhibited statistically significant associations between several explanatory variables and being overweight or obese. Notably, male children exhibited a higher likelihood of being OW/OB, with 1.26 times increased adjusted odds (95% CI: 1.20 to 1.33) compared to female children. In addition, for every yearly increase in a child’s age, the odds of being overweight or obese increased by 1.10 times (95% CI: 1.09 to 1.10). When broken down by ethnicity, compared to Indigenous children, the Afroecuadorian ethnicity presented a slightly elevated but not statistically significant odds of 1.12 (95% CI: 0.99 to 1.26), while Mestizo children showed 1.14 times increased odds (95% CI: 1.04 to 1.25). White children and those from Montubio (mixed ethnic background of coastal Ecuador) or other ethnicities did not demonstrate statistically significant differences in this model. When considering economic quintiles by income, children in the 2nd quintile demonstrated 1.17 times higher odds (95% CI: 1.07 to 1.31), those in the 3rd quintile showed 1.33 times (95% CI: 1.11 to 1.59), in the 4th quintile it was 1.39 times (95% CI: 1.18 to 1.65), and in the 5th quintile, the odds were 1.39 times higher (95% CI: 1.29 to 1.51) compared to those in the 1st quintile. An increase in the number of household members corresponded to a slight reduction in odds by 0.93 times for each additional person (95% CI: 0.91 to 0.95). Moreover, children with inadequate disposal of excreta exhibited 0.82 times lower odds of being overweight or obese (95% CI: 0.76 to 0.90). Similarly, regular physical activity was associated with reduced odds, at 0.79 times (95% CI: 0.75 to 0.82). Interestingly, children from families that recognized and used processed food labels exhibited a higher likelihood of being overweight or obese, with an adjusted odds ratio (aOR) of 1.14 (95% CI: 1.02 to 1.26). Conversely, the consumption of food provided by schools was linked with a non-significant reduction in the risk of overweight or obesity, with an aOR of 0.93 (95% CI: 0.82 to 1.06).

**Table 2.**
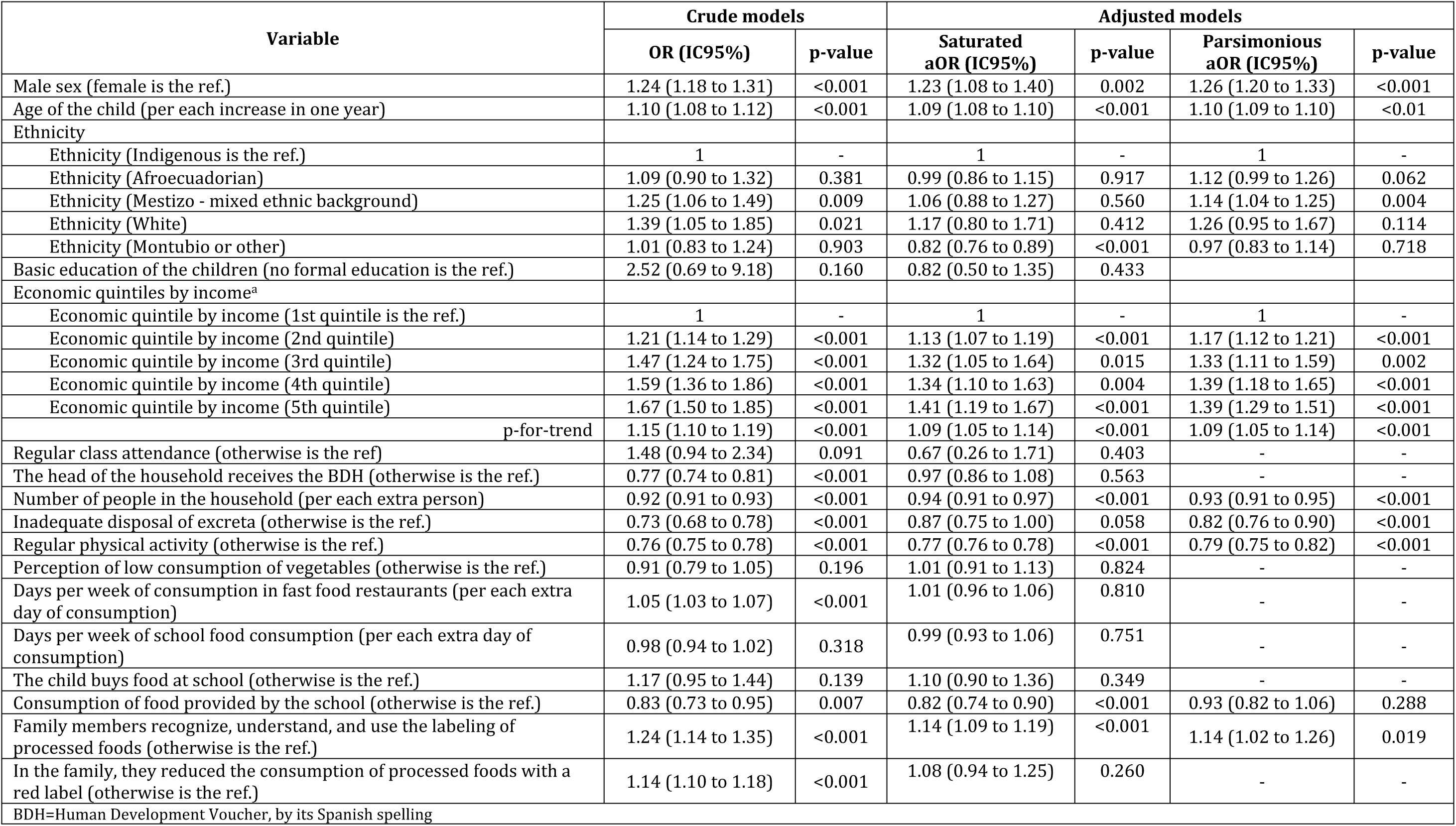
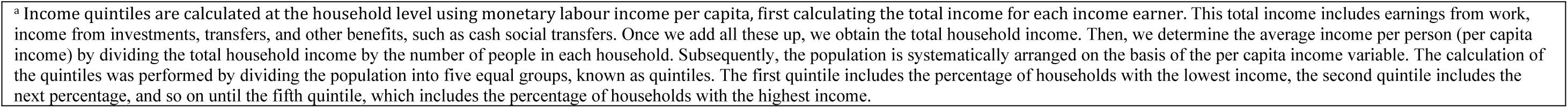
Crude and adjusted Odds Ratios of overweight or obesity from each explanatory variable using multilevel and logistic regression models.

| Variable | Crude models |  | Adjusted models |  |  |  |
| --- | --- | --- | --- | --- | --- | --- |
|  | OR (IC95%) | p-value | Saturated aOR (IC95%) | p-value | Parsimonious aOR (IC95%) | p-value |
| Male sex (female is the ref.) | 1.24 (1.18 to 1.31) | <0.001 | 1.23 (1.08 to 1.40) | 0.002 | 1.26 (1.20 to 1.33) | <0.001 |
| Age of the child (per each increase in one year) | 1.10 (1.08 to 1.12) | <0.001 | 1.09 (1.08 to 1.10) | <0.001 | 1.10 (1.09 to 1.10) | <0.01 |
| Ethnicity |  |  |  |  |  |  |
| Ethnicity (Indigenous is the ref.) | 1 | - | 1 | - | 1 | - |
| Ethnicity (Afroecuadorian) | 1.09 (0.90 to 1.32) | 0.381 | 0.99 (0.86 to 1.15) | 0.917 | 1.12 (0.99 to 1.26) | 0.062 |
| Ethnicity (Mestizo - mixed ethnic background) | 1.25 (1.06 to 1.49) | 0.009 | 1.06 (0.88 to 1.27) | 0.560 | 1.14 (1.04 to 1.25) | 0.004 |
| Ethnicity (White) | 1.39 (1.05 to 1.85) | 0.021 | 1.17 (0.80 to 1.71) | 0.412 | 1.26 (0.95 to 1.67) | 0.114 |
| Ethnicity (Montubio or other) | 1.01 (0.83 to 1.24) | 0.903 | 0.82 (0.76 to 0.89) | <0.001 | 0.97 (0.83 to 1.14) | 0.718 |
| Basic education of the children (no formal education is the ref.) | 2.52 (0.69 to 9.18) | 0.160 | 0.82 (0.50 to 1.35) | 0.433 |  |  |
| Economic quintiles by income<sup>a</sup> |  |  |  |  |  |  |
| Economic quintile by income (1st quintile is the ref.) | 1 | - | 1 | - | 1 | - |
| Economic quintile by income (2nd quintile) | 1.21 (1.14 to 1.29) | <0.001 | 1.13 (1.07 to 1.19) | <0.001 | 1.17 (1.12 to 1.21) | <0.001 |
| Economic quintile by income (3rd quintile) | 1.47 (1.24 to 1.75) | <0.001 | 1.32 (1.05 to 1.64) | 0.015 | 1.33 (1.11 to 1.59) | 0.002 |
| Economic quintile by income (4th quintile) | 1.59 (1.36 to 1.86) | <0.001 | 1.34 (1.10 to 1.63) | 0.004 | 1.39 (1.18 to 1.65) | <0.001 |
| Economic quintile by income (5th quintile) | 1.67 (1.50 to 1.85) | <0.001 | 1.41 (1.19 to 1.67) | <0.001 | 1.39 (1.29 to 1.51) | <0.001 |
| p-for-trend | 1.15 (1.10 to 1.19) | <0.001 | 1.09 (1.05 to 1.14) | <0.001 | 1.09 (1.05 to 1.14) | <0.001 |
| Regular class attendance (otherwise is the ref) | 1.48 (0.94 to 2.34) | 0.091 | 0.67 (0.26 to 1.71) | 0.403 | - | - |
| The head of the household receives the BDH (otherwise is the ref.) | 0.77 (0.74 to 0.81) | <0.001 | 0.97 (0.86 to 1.08) | 0.563 | - | - |
| Number of people in the household (per each extra person) | 0.92 (0.91 to 0.93) | <0.001 | 0.94 (0.91 to 0.97) | <0.001 | 0.93 (0.91 to 0.95) | <0.001 |
| Inadequate disposal of excreta (otherwise is the ref.) | 0.73 (0.68 to 0.78) | <0.001 | 0.87 (0.75 to 1.00) | 0.058 | 0.82 (0.76 to 0.90) | <0.001 |
| Regular physical activity (otherwise is the ref.) | 0.76 (0.75 to 0.78) | <0.001 | 0.77 (0.76 to 0.78) | <0.001 | 0.79 (0.75 to 0.82) | <0.001 |
| Perception of low consumption of vegetables (otherwise is the ref.) | 0.91 (0.79 to 1.05) | 0.196 | 1.01 (0.91 to 1.13) | 0.824 | - | - |
| Days per week of consumption in fast food restaurants (per each extra day of consumption) | 1.05 (1.03 to 1.07) | <0.001 | 1.01 (0.96 to 1.06) | 0.810 | - | - |
| Days per week of school food consumption (per each extra day of consumption) | 0.98 (0.94 to 1.02) | 0.318 | 0.99 (0.93 to 1.06) | 0.751 | - | - |
| The child buys food at school (otherwise is the ref.) | 1.17 (0.95 to 1.44) | 0.139 | 1.10 (0.90 to 1.36) | 0.349 | - | - |
| Consumption of food provided by the school (otherwise is the ref.) | 0.83 (0.73 to 0.95) | 0.007 | 0.82 (0.74 to 0.90) | <0.001 | 0.93 (0.82 to 1.06) | 0.288 |
| Family members recognize, understand, and use the labeling of processed foods (otherwise is the ref.) | 1.24 (1.14 to 1.35) | <0.001 | 1.14 (1.09 to 1.19) | <0.001 | 1.14 (1.02 to 1.26) | 0.019 |
| In the family, they reduced the consumption of processed foods with a red label (otherwise is the ref.) | 1.14 (1.10 to 1.18) | <0.001 | 1.08 (0.94 to 1.25) | 0.260 | - | - |
| BDH=Human Development Voucher, by its Spanish spelling |  |  |  |  |  |  |

<sup>a</sup> Income quintiles are calculated at the household level using monetary labour income per capita, first calculating the total income for each income earner. This total income includes earnings from work, income from investments, transfers, and other benefits, such as cash social transfers. Once we add all these up, we obtain the total household income. Then, we determine the average income per person (per capita income) by dividing the total household income by the number of people in each household. Subsequently, the population is systematically arranged on the basis of the per capita income variable. The calculation of the quintiles was performed by dividing the population into five equal groups, known as quintiles. The first quintile includes the percentage of households with the lowest income, the second quintile includes the next percentage, and so on until the fifth quintile, which includes the percentage of households with the highest income.

In analyzing the determinants of overweight and obesity in children, notable differences emerged between genders when running the final parsimonious model (Table 3). For each incremental year in age, a significant association with overweight or obesity was noted in both genders, with the odds of being overweight or obese increasing by an adjusted odds ratio (aOR) of 1.09 (95% CI: 1.07 to 1.11) for women and 1.10 (95% CI: 1.09 to 1.11) for men. Among the ethnicities, White ethnicity was associated with the highest risk in women, with an aOR of 1.57 (95% CI: 1.34 to 1.83). With respect to economic quintiles, women in the 5th quintile exhibited the greatest risk, with aOR of 1.38 (95% CI: 1.13 to 1.70). In relation to other determining factors, it was observed that inadequate disposal of excreta had significant associations with overweight or obesity in both genders. For women, the risk was reduced with an adjusted odds ratio (aOR) of 0.86 (95% CI: 0.80 to 0.92), whereas for men, the reduction in risk was slightly more pronounced with an aOR of 0.81 (95% CI: 0.74 to 0.89). Regular physical activity appeared to be protective against overweight or obesity. Women who engaged in regular physical activity presented a reduced risk, as indicated by an aOR of 0.84 (95% CI: 0.80 to 0.88), whereas men benefitted slightly more from such activity, displaying an aOR of 0.76 (95% CI: 0.70 to 0.83). Notably, in households where processed food labeling was recognized, understood, and utilized, the risk of overweight or obesity increased in both women and men. This association, for women, was statistically significant with an aOR of 1.16 (95% CI: 1.09 to 1.24), and for men, it was not significant (aOR= 1.11; 95% CI: 0.95 to 1.29).

**Table 3.** Adjusted Odds Ratios of overweight or obesity from each explanatory variable using the parsimonious logistic regression model of Table 2 between women and men.

| Variable | Parsimonious model in women<br>aOR (IC95%) | p-value | Parsimonious model in men<br>aOR (IC95%) | p-value |
| --- | --- | --- | --- | --- |
| Age of the child (per each increase in one year) | 1.09 (1.07 to 1.11) | <0.001 | 1.10 (1.09 to 1.11) | <0.001 |
| Ethnicity |  |  |  |  |
| Ethnicity (Indigenous is the ref.) | 1 | - | 1 | - |
| Ethnicity (Afroecuadorian) | 1.19 (0.82 to 1.72) | 0.374 | 1.07 (0.93 to 1.23) | 0.325 |
| Ethnicity (Mestizo – mixed ethnic background) | 1.32 (1.09 to 1.60) | 0.005 | 1.01 (0.97 to 1.06) | 0.653 |
| Ethnicity (White) | 1.57 (1.34 to 1.83) | <0.001 | 1.05 (0.64 to 1.70) | 0.858 |
| Ethnicity (Montubio or other) | 0.94 (0.67 to 1.32) | 0.716 | 0.99 (0.97 to 1.00) | 0.196 |
| Economic quintiles by income<sup>a</sup> |  |  |  |  |
| Economic quintile by income (1st quintile is the ref.) | 1 | - | 1 | - |
| Economic quintile by income (2nd quintile) | 1.13 (1.10 to 1.17) | <0.001 | 1.19 (1.14 to 1.24) | <0.001 |
| Economic quintile by income (3rd quintile) | 1.27 (0.88 to 1.83) | 0.203 | 1.38 (1.32 to 1.44) | <0.001 |
| Economic quintile by income (4th quintile) | 1.31 (1.07 to 1.60) | 0.009 | 1.48 (1.26 to 1.73) | <0.001 |
| Economic quintile by income (5th quintile) | 1.38 (1.13 to 1.70) | 0.002 | 1.40 (1.33 to 1.48) | <0.001 |
| p-for-trend | 1.09 (1.01 to 1.17) | 0.021 | 1.10 (1.09 to 1.12) | <0.001 |
| Number of people in the household (per each extra person) | 0.92 (0.90 to 0.95) | <0.001 | 0.93 (0.88 to 1.00) | 0.041 |
| Inadequate disposal of excreta (otherwise is the ref.) | 0.86 (0.80 to 0.92) | <0.001 | 0.81 (0.74 to 0.89) | <0.001 |
| Regular physical activity (otherwise is the ref.) | 0.84 (0.80 to 0.88) | <0.001 | 0.76 (0.69 to 0.83) | <0.001 |
| Consumption of food provided by the school (otherwise is the ref.) | 1.01 (0.98 to 1.05) | 0.499 | 0.86 (0.69 to 1.09) | <0.001 |
| Family members recognize, understand, and use the labeling of processed foods (otherwise is the ref.) | 1.16 (1.09 to 1.24) | <0.001 | 1.11 (0.95 to 1.29) | 0.179 |

BDH=Human Development Voucher, by its Spanish spelling
<sup>a</sup> Income quintiles are calculated at the household level using monetary labour income per capita, first calculating the total income for each income earner. This total income includes earnings from work, income from investments, transfers, and other benefits, such as cash social transfers. Once we add all these up, we obtain the total household income. Then, we determine the average income per person (per capita income) by dividing the total household income by the number of people in each household. Subsequently, the population is systematically arranged on the basis of the per capita income variable. The calculation of the quintiles was performed by dividing the population into five equal groups, known as quintiles. The first quintile includes the percentage of households with the lowest income, the second quintile includes the next percentage, and so on until the fifth quintile, which includes the percentage of households with the highest income.

After conducting an analysis excluding children in the highest quintile, children with parents receiving the BDH, and those residing in households with a larger number of individuals, the observed associations maintained similar trends. However, the relationships were less statistically significant, as detailed in S3 Table.

## Discussion

### Principle Findings

Increasing age, male gender, mestizo (mixed ethnic background) ethnicity, higher economic quintiles, inadequate disposal of excreta, and lack of physical activity are factors associated with a higher likelihood of overweight or obesity in children aged 5 to 11 years in Ecuador. The impact of consuming school-provided meals was inconclusive. Children from families with a higher number of individuals in the household and with families that recognize and use processed food labels exhibited a higher likelihood of being overweight or obese.

### Comparison with the Literature

The prevalence of OW/OB in school children places the country in the eighth position in the Americas, following countries such as Mexico, Chile, Panama, and the United States, and surpassing the prevalences in Colombia and Peru [22]. International data reported in 2016 ranked the country 15th in the same region [23]. This highlights the drastic increase in prevalence in the absence of effective public policies, compared with other countries that have taken strong measures against childhood overweight and obesity [24,25].

The study reveals that children who purchase food at school are at a greater risk of being overweight or obese than those who consume meals provided by the school, but this difference is not statistically significant. Despite that, it is important to mention that prior research from the ENSANUT 2018 survey, indicated a link between school foods, particularly those sold in stores (73%), and elevated BMI [26]. A plausible explanation is that 60% of these stores provide unhealthy products labeled with a “red traffic light”. The lack of significant results may stem from the Ecuadorian school feeding program, which relies on processed and packaged products mandated to bear nutritional labeling (traffic light system) [1]. These products contain high levels of sugar, salt, and fats similar to the ultra-processed items distributed by food industries in formal markets.

The study underscores the crucial role of the school food environment in addressing childhood obesity. Despite robust evidence[27], many Latin American schools, including those in Ecuador[28]. lack effective public health policies. Practical recommendations include prohibiting the advertisement and sale of processed foods in and around schools, imposing higher taxes on sugar-rich, ultra-processed products, and promoting the consumption of fresh fruits and vegetables, along with education on food labeling[29].

In a broader epidemiological context, the gradual increase in the prevalence of OW/OB in the population of this study, along with a population of adults who have been overweight and obese for decades, coupled with a distribution of excess malnutrition across all socioeconomic strata in both children and adults [9], places Ecuador in a second stage of transitioning to obesity. It is noteworthy that the richest quintile of households has a lower prevalence of OW/OB than the quintile immediately below, which could indicate that Ecuador is entering the third stage of transitioning to obesity, where excess malnutrition decreases in wealthier households and becomes concentrated in socioeconomically vulnerable households [5]. Additionally, the observation that children residing in homes with inadequate excreta disposal are less likely to be OW/OB underscores the presence of a socioeconomic gradient in obesity and overweight prevalence in Ecuador. This characteristic is compounded by the fact that the country has not overcome malnutrition in early childhood [9], further complicating the issue of overall malnutrition, resulting in a country experiencing a double burden of malnutrition (malnutrition due to deficiency and excess).

The prevalence patterns of OW/OB in Ecuadorian school children are similar to those in other low and middle-income countries in the region. Similar to a study of school children in Mexico, there is a higher prevalence of OW/OB in boys than in girls, in the non-indigenous population, and in higher-income households, and OW/OB also increases with age [30,31]. However, the results of this study differ from patterns seen in high-income countries in the region such as Chile, where OW/OB is more prevalent in socioeconomically vulnerable families [32]. It is challenging to compare the determinants found in this study with those in other countries in the region because of to the scarcity of published studies [33].

The fact that children from households self-identified as mestizo have higher rates of OW/OB may be explained by the fact that these children tend to live in urban environments where they are more exposed to obesogenic environments such as advertisements for sugary beverages, ultra-processed foods, and junk food. This would also explain why indigenous children are protected from obesity and overweight, in addition to socioeconomic vulnerabilities in this population, along with household size, which is more strongly linked to malnutrition due to deficiency rather than excess in the country [15,34]. Therefore, further research is needed on the circumstances of social vulnerability and food insecurity in households of school children with OW/OB to explain why certain negative socioeconomic factors seem to protect against OW/OB.

Physical activity among schoolchildren acts as a protective factor against OW/OB [35,36]. However, the percentage of children in the sample who engage in regular physical activity is very low, which aligns with previously published findings [37]. There is limited research linking physical activity to excess malnutrition in this group. Therefore, further research on this topic is necessary.

Regarding gender differences, our findings support the current understanding of gender-based dietary segregation, both in Ecuador and globally. In Ecuador, prevailing dietary customs, shaped by a patriarchal and sexist societal framework, often prioritize feeding boys. This may partly account for why physical activity appears to benefit boys more significantly in reducing the likelihood of being overweight or obese, as compared to girls, where such activities do not seem to diminish the probability of overweight or obesity as effectively [38]. This is exacerbated by the inclination to maintain purely aesthetic standards, which results in food restrictions for girls, especially as they enter adolescence [39]. Additionally, there are existing and differential unhealthy exposures based on socioeconomic strata [40].

Given Ecuador’s extensive history of childhood chronic malnutrition, a pivotal risk factor for overweight, obesity, and non-communicable diseases, coupled with the severe impacts of climate change linked to environmental factors, it is imperative to focus research and public policies on understanding the global syndemic of malnutrition. This approach should incorporate triple-duty actions, considering the distinctive aspects of diverse life stages. [41].

Our findings suggest that public health policies should place greater focus on improving the quality of available foods within schools to mitigate the risks of childhood overweight and obesity.

In the context of Ecuador and the wider region, our study underscores the criticality of scrutinizing policy missteps that deregulate the marketing of unhealthy foods and other harmful products, thereby hindering the enhancement of food environments. Remarkably, the President of Ecuador enacted Presidential Decree No. 645 on January 10, 2023, which aims to slash taxes on known health hazards, such as alcohol, tobacco, sugary beverages, and firearms, in stark contrast to advancing public health policies [16]. This decree is poised to adversely affect collective health. This underscores a significant lacuna in public health strategies, potentially exacerbating the obesity and overweight crisis among school children. This situation urgently demands the attention of authorities and policymakers to realign regulations with public health principles, ensuring that economic interests and conflicts of interest do not undermine the development of sound public health policies [42,43]. In this context, it becomes imperative for Ecuadorian authorities to implement and strengthen comprehensive policies that address both the quality of food offered within schools and those sold in their vicinity.

The congruence between the primary and sensitivity analyses bolsters the robustness of our findings. Although the consumption of food purchased at school and food provided by the school did not attain statistical significance in the model adjusted for expansion factors, the overall pattern of results remained consistent. Importantly, the existing scientific literature supports our findings concerning these variables. It is also crucial to emphasize that there are additional food environments associated with formal markets and marketing targeted at children, which were not considered in this study but significantly contribute to childhood OW/OB. Consequently, it is imperative that national health and nutrition surveys not only possess sensitivity to growth retardation but also to excess malnutrition. While the study delves into various sociodemographic factors, some potentially relevant variables, such as cultural practices or parental education, are not thoroughly examined. It is worth mentioning that not all information from different forms can be cross-referenced for analysis because of to variations in the design and objectives of the national survey. We believe that our findings necessitate authorities to contemplate public policies aimed at reducing the burden of OW/OB from an early age, thereby diminishing future disabilities and deaths, which evidently result in heavier economic costs and social burdens.

## Conclusions

Increasing age, male gender, mestizo (mixed ethnic background) ethnicity, higher economic quintiles, inadequate disposal of excreta, and lack of physical activity are factors associated with a higher likelihood of overweight or obesity in children aged 5 to 11 years in Ecuador. The impact of consuming school-provided meals was inconclusive. Children from families with a higher number of individuals in the household and with families that recognize and use processed food labels exhibited a higher likelihood of being overweight or obese. It is imperative for the Ecuadorian government to implement public policy actions aimed at safeguarding the right to health of school-age children, addressing both social protection within households and evidence-based dietary policies for promoting healthy school food environments.

## Data Availability

All relevant data are within the manuscript and its Supporting Information files.

## Acknowledgements

We extend our sincere gratitude to David Grijalva, an economics student from the Faculty of Economics at the Pontifical Catholic University of Ecuador, for his invaluable assistance in the preparation of the tables for this manuscript. Additionally, we would like to thank Natali Mendoza and Margoth Herrera from the Department of Sociodemographic Statistics at the National Institute of Statistics and Censuses for their specific technical guidance on the online database.

## Author Contribution

**Conceptualization:** Betzabé Tello, Iván Dueñas-Espín, José Ocaña, Paúl García-Zambrano, Betsabé Enríquez-Moreira.

**Data curation:** Betzabé Tello, Iván Dueñas-Espín, José Ocaña

**Formal analysis:** Iván Dueñas-Espín

**Funding acquisition:** Betzabé Tello

**Investigation:** Betzabé Tello, Iván Dueñas-Espín

**Methodology:** Betzabé Tello, Iván Dueñas-Espín, José Ocaña

**Project administration:** Betzabé Tello

**Resources:** Betzabé Tello, Iván Dueñas-Espín, José Ocaña

**Software:** Iván Dueñas-Espín, José Ocaña

**Supervision:** Betzabé Tello

**Validation:** Betzabé Tello, Iván Dueñas-Espín, José Ocaña, Paúl García-Zambrano, Betsabé Enríquez-Moreira.

**Visualization:** Betzabé Tello, Iván Dueñas-Espín

**Writing – original draft:** Betzabé Tello, Iván Dueñas-Espín, José Ocaña

**Writing – review & editing:** Betzabé Tello, Iván Dueñas-Espín, José Ocaña, Paúl García-Zambrano, Betsabé Enríquez-Moreira.

## Notes

### Competing Interest Statement

The authors have declared no competing interest.

### Funding Statement

The author(s) received no specific funding for this work.

## References

1. Popkin BM, Adair LS, Ng SW. Global nutrition transition and the pandemic of obesity in developing countries. Nutr Rev. 2012;70: 3–21. doi:10.1111/j.1753-4887.2011.00456.x

2. Popkin BM. Global nutrition dynamics: the world is shifting rapidly toward a diet linked with noncommunicable diseases1–3. Am J Clin Nutr. 2006;84: 289–298. doi:10.1093/ajcn/84.1.289

3. Monteiro CA, Moubarac JC, Cannon G, Ng SW, Popkin B. Ultra-processed products are becoming dominant in the global food system. Obes Rev. 2013;14: 21–28. doi:10.1111/obr.12107

4. Di Cesare M, Sorić M, Bovet P, Miranda JJ, Bhutta Z, Stevens GA, et al. The epidemiological burden of obesity in childhood: A worldwide epidemic requiring urgent action. BMC Med. 2019;17. doi:10.1186/s12916-019-1449-8

5. Jaacks LM, Vandevijvere S, Pan A, McGowan CJ, Wallace C, Imamura F, et al. The obesity transition: stages of the global epidemic. The Lancet Diabetes and Endocrinology. Europe PMC Funders; 2019. pp. 231–240. doi:10.1016/S2213-8587(19)30026-9

6. Umer A, Kelley GA, Cottrell LE, Giacobbi P, Innes KE, Lilly CL. Childhood obesity and adult cardiovascular disease risk factors: A systematic review with meta-analysis. BMC Public Health. 2017;17. doi:10.1186/s12889-017-4691-z

7. Weihrauch-Blüher S, Schwarz P, Klusmann JH. Childhood obesity: increased risk for cardiometabolic disease and cancer in adulthood. Metabolism. 2019;92: 147–152. doi:10.1016/j.metabol.2018.12.001

8. Callo Quinte G, Barros F, Gigante DP, De Oliveira IO, Dos Santos Motta JV, Horta BL. Overweight trajectory and cardio metabolic risk factors in young adults. BMC Pediatr. 2019;19. doi:10.1186/s12887-019-1445-3

9. Cando F, Diego M, Pozo M. Reportes de la ENSANUT 2018. Volumen No. 3 Antropometría. Inst Nac Estadística y Censos. Quito; 2022. Available: chrome-e/https://www.ecuadorencifras.gob.ec/documentos/web-inec/Bibliotecas/Libros/Reportes/Reportes_ENSANUT_Vol3_Antropometria.pdf

10. Jebeile H, Kelly AS, O’Malley G, Baur LA. Obesity in children and adolescents: epidemiology, causes, assessment, and management. Lancet Diabetes Endocrinol. 2022;10: 351–365. doi:10.1016/S2213-8587(22)00047-X

11. Avelar Rodriguez D, Toro Monjaraz EM, Ignorosa Arellano KR, Ramirez Mayans J. Childhood obesity in Mexico: Social determinants of health and other risk factors. BMJ Case Rep. 2018;2018. doi:10.1136/bcr-2017-223862

12. Corvalán C, Garmendia ML, Jones-Smith J, Lutter CK, Miranda JJ, Pedraza LS, et al. Nutrition status of children in Latin America. Obes Rev. 2017;18: 7–18. doi:10.1111/obr.12571

13. Freire WB, Silva-Jaramillo KM, Ramirez-Luzuriaga MJ, Belmont P, Waters WF. The double burden of undernutrition and excess body weight in Ecuador. Am J Clin Nutr. 2014;100: 1636S–1643S. doi:10.3945/ajcn.114.083766

14. Abril V, Manuel-Y-Keenoy B, Solà R, García JL, Nessier C, Rojas R, et al. Prevalence of overweight and obesity among 6- to 9-year-old schoolchildren in cuenca, ecuador: Relationship with physical activity, poverty, and eating habits. Food Nutr Bull. 2013;34: 388–400. doi:10.1177/156482651303400404

15. Walrod J, Seccareccia E, Sarmiento I, Pimentel JP, Misra S, Morales J, et al. Community factors associated with stunting, overweight and food insecurity: A community-based mixed-method study in four Andean indigenous communities in Ecuador. BMJ Open. 2018;8. doi:10.1136/bmjopen-2017-020760

16. Presidencia de la República del Ecuador. Decreto Ejecutivo 645. 2023.

17. Von Elm E, Altman DG, Egger M, Pocock SJ, Gøtzsche PC, Vandenbrouckef JP. The Strengthening the Reporting of Observational Studies in Epidemiology (STROBE) Statement: Guidelines for reporting observational studies. Bull World Health Organ. 2007;85: 867–872. doi:10.2471/BLT.07.045120

18. Mendoza N, Ocaña N, Guano D, Núñez J, Valdivieso K. Documento Metodológico de la Encuesta Nacional de Salud y Nutrición (ENSANUT). Inst Nac Estadística y Censos. Quito - Ecuador; 2019. Available: https://www.ecuadorencifras.gob.ec/documentos/web-inec/Estadisticas_Sociales/ENSANUT/ENSANUT_2018/MetodologiaENSANUT2018.pdf

19. Ministerio de Salud Pública del Ecuador. Reglamento de Etiquetado de Alimentos Procesados para Consumo Humano. Registro Oficial Suplemento Quito - Ecuador: https://www.controlsanitario.gob.ec/wp-content/uploads/downloads/2016/12/Reglamento-de-Etiquetado-de-Alimentos-procesados-para-consumo-humano.pdf; 2014. Available: https://www.controlsanitario.gob.ec/wp-content/uploads/downloads/2014/08/REGLAMENTO-SANITARIO-DE-ETIQUETADO-DE-ALIMENTOS-PROCESADOS-PARA-EL-CONSUMO-HUMANO-junio-2014.pdf

20. World Health Organization. Growth Reference data for 5-19 years. Available: https://www.who.int/tools/growth-reference-data-for-5to19-years/indicators/bmi-for-age

21. Harrell FE. Regression Modeling Strategies with applications to linear models, logistic and ordinal regression and survival analysis. Second. Springer International Publishing, editor. Journal of Statistical Software. Switzerland; 2015. doi:10.1007/978-3-319-19425-7

22. World Obesity Federation. Prevalence of child overweight, including obesity (%). In: Global Obesity Observatory [Internet]. 2023 [cited 26 Sep 2023]. Available: https://data.worldobesity.org/tables/prevalence-of-child-overweight-including-obesity-3/?regionid=3&msr=msr&breakdown=c

23. Our world in data. Share of children and adolescents that are overweight or obese, 1975 to 2016. 2022 [cited 26 Sep 2023]. Available: https://ourworldindata.org/grapher/child-adolescent-obesity?tab=chart

24. Duran AC, Mialon M, Crosbie E, Jensen ML, Harris JL, Batis C, et al. Food environment solutions for childhood obesity in Latin America and among Latinos living in the United States. Obes Rev. 2021;22. doi:10.1111/obr.13237

25. Pérez-Escamilla R, Vilar-Compte M, Rhodes E, Sarmiento OL, Corvalan C, Sturke R, et al. Implementation of childhood obesity prevention and control policies in the United States and Latin America: Lessons for cross-border research and practice. Obes Rev. 2021;22. doi:10.1111/obr.13247

26. Weigel MM, Armijos RX. The Ecuadorian School Food Environment: Association With Healthy and Unhealthy Food and Beverage Consumption and BMI. Food Nutr Bull. 2022;43: 439–464. doi:10.1177/03795721221116447

27. Hawkes C, Smith TG, Jewell J, Wardle J, Hammond RA, Friel S, et al. Smart food policies for obesity prevention. The Lancet. Elsevier; 2015. pp. 2410–2421. doi:10.1016/S0140-6736(14)61745-1

28. Ministerio de Salud Pública del Ecuador, Ministerio de Educación. Reglamento de bares escolares del Sistema Nacional de Educación. Ecuador; 2020 pp. 1–1. Available: https://www.controlsanitario.gob.ec/wp-content/plugins/download-monitor/download.php?id=9565&force=1

29. Temple NJ. A Proposed Strategy against Obesity: How Government Policy Can Counter the Obesogenic Environment. Nutrients. 2023;15. doi:10.3390/nu15132910

30. del Monte Vega MY, Ávila Curiel A, Ávila Arcos MA, Galindo Gómez C, Shamah Levy T. Overweight and obesity in the Mexican school-age population from 2015 to 2019. Nutr Hosp. 2022;39: 1076–1085. doi:10.20960/nh.04028

31. Shamah-Levy T, Cuevas-Nasu L, Humarán IMG, Morales-Ruán C, Valenzuela-Bravo DG, Gaona-Pineda EB, et al. Prevalencia y predisposición a la obesidad en una muestra nacional de niños y adolescentes en México. Salud Publica Mex. 2020;62: 725–733. doi:10.21149/11552

32. Amigo C. H, Bustos P, Erazo M, Cumsille P, Silva C. Factores determinantes del exceso de peso en escolares: Un estudio multinivel. Rev Med Chil. 2007;135: 1510–1518. doi:10.4067/s0034-98872007001200002

33. Allender S, Owen B, Kuhlberg J, Lowe J, Nagorcka-Smith P, Whelan J, et al. A community based systems diagram of obesity causes. PLoS One. 2015;10: 1–12. doi:10.1371/journal.pone.0129683

34. Rivadeneira MF, Moncayo AL, Cóndor JD, Tello B, Buitrón J, Astudillo F, et al. High prevalence of chronic malnutrition in indigenous children under 5 years of age in Chimborazo-Ecuador: multicausal analysis of its determinants. BMC Public Health. 2022;22. doi:10.1186/s12889-022-14327-x

35. Ip P, Ho FKW, Louie LHT, Chung TWH, Cheung YF, Lee SL, et al. Childhood Obesity and Physical Activity-Friendly School Environments. J Pediatr. 2017;191: 110–116. doi:10.1016/j.jpeds.2017.08.017

36. Yuksel HS, Şahin FN, Maksimovic N, Drid P, Bianco A. School-based intervention programs for preventing obesity and promoting physical activity and fitness: A systematic review. Int J Environ Res Public Health. 2020;17. doi:10.3390/ijerph17010347

37. Andrade S, Ochoa-Avilés A, Freire W, Romero-Sandoval N, Orellana D, Contreras T, et al. Results from ecuador’s 2018 report card on physical activity for children and youth. J Phys Act Heal. 2018;15: S344–S346. doi:10.1123/JPAH.2018-0536

38. Flores J. Soberanía alimentaria y mujeres. Instituto de Estudios Ecuatorianos, editor. IEE Soberanía Alimentaria y mujeres. Quito; 2013. Available: https://www.iee.org.ec/publicaciones/sociedad-alternativa/soberania-alimentaria-y-mujeres.html

39. Wang S, Sun Q, Zhai L, Bai Y, Wei W, Jia L. The prevalence of depression and anxiety symptoms among overweight/obese and non-overweight/non-obese children/adolescents in China: A systematic review and meta-analysis. Int J Environ Res Public Health. 2019;16. doi:10.3390/ijerph16030340

40. Isong IA, Rao SR, Bind MA, Avendaño M, Kawachi I, Richmond TK. Racial and ethnic disparities in early childhood obesity. Pediatrics. 2018;141. doi:10.1542/peds.2017-0865

41. Venegas Hargous C, Strugnell C, Allender S, Orellana L, Corvalan C, Bell C. Double- and triple-duty actions in childhood for addressing the global syndemic of obesity, undernutrition, and climate change: A scoping review. Obes Rev. 2023;24. doi:10.1111/obr.13555

42. Rogers NT, Cummins S, Forde H, Jones CP, Mytton O, Rutter H, et al. Associations between trajectories of obesity prevalence in English primary school children and the UK soft drinks industry levy: An interrupted time series analysis of surveillance data. PLoS Med. 2023;20: 1–18. doi:10.1371/journal.pmed.1004160

43. Carriedo A, Cairney P, Barquera S, Hawkins B. Policy networks and competing interests in the development of the Mexican sugar-sweetened beverages tax. BMJ Glob Heal. 2023;8: 1–15. doi:10.1136/bmjgh-2023-012125

